# The Use of Procalcitonin as an Antimicrobial Stewardship Tool and a Predictor of Disease Severity in COVID-19

**DOI:** 10.1101/2021.01.14.21249853

**Authors:** George P Drewett, Olivia C Smibert, Natasha E Holmes, Jason A Trubiano

## Abstract

In our study, procalcitonin was associated with both antibiotic use and duration in patients with COVID-19, as well as established biochemical markers of COVID-19 disease severity and oxygen requirement, suggesting a potential role for procalcitonin in COVID-19 antimicrobial stewardship.

## Introduction

Prior to COVID-19, serum procalcitonin (PCT)-based antimicrobial stewardship (AMS) algorithms have been shown to be effective at differentiating between bacterial and non-bacterial respiratory tract infection, leading to improved mortality, less antibiotic use, and decreased risk of antibiotic side effects(3). In COVID-19, initial reports of the utility of PCT were from hospitalised patients where PCT was found to correlate with disease severity, longer intensive care unit (ICU) stay, and inpatient mortality, along with a range of other biochemical markers (4-6). However, PCT would have unique value if its measurement at admission for COVID-19 illness or at time of clinical deterioration could be an important discriminator between bacterial co-infection versus non-infectious cause, allowing for improved AMS (7), as there is widespread use of empiric antibiotics globally in hospitalised patients with COVID-19, despite low rates of microbiologically proven bacterial infection (1, 2).

We investigated whether PCT was associated with commencement of antibiotic therapy. Additionally, we examined if PCT was associated with duration of antibiotic therapy, intravenous-to-oral antibiotic switch, and other clinical and biochemical markers of COVID-19 disease severity.

## Methods

A single-centre, prospective, observational cohort study of patients with COVID-19 admitted to Austin Health (Melbourne, Australia) was undertaken. All patients were admitted to a specialized, multidisciplinary unit coordinated by Infectious Diseases Physicians. PCT was measured at time of ICU admission, or at clinician discretion outside the ICU. Patients were stratified based on their initial PCT measurement into normal (<0.07mcg/L), medium (0.07mcg/L – 0.5mcg/L) and high (>0.5mcg/L) groups, based on consensus guidelines and recently published data in a patient group with COVID-19 (5, 8). Demographic, clinical and laboratory data were also collected. Statistical analysis was performed using Stata MP 16.1 (StataCorp, College Station, TX). Chi square and rank sum tests were used for univariate analysis according to PCT strata. Given the limited sample size, multivariable analysis was not performed.

## Results

166 patients were admitted with COVID-19 at the Austin Hospital March and September 2020. Of these, 55 had at least one PCT measurement during their admission (Table 1). Most patients had PCT measured within the first day of admission (median (days): 1. IQR 0,3). Blood cultures were taken in 42/55 patients, with three positive results (4.7%).

**Table 1:** Demographics, laboratory parameters, treatment parameters and outcomes for normal PCT, medium PCT and high PCT groups

|  | Normal PCT group<br>(PCT <0.07mcg/L) | Medium PCT group<br>(PCT 0.07-0.5mcg/L) | High PCT group<br>(PCT >0.5mcg/L) | p-value |
| --- | --- | --- | --- | --- |
| N | 15 | 27 | 13 |  |
| **Demographics** |  |  |  |  |
| Age in years, median (IQR) | 61 (34, 82) (n=15) | 66 (59, 76) (n=27) | 62 (56, 79) (n=13) | 0.81 |
| Female sex | 9 (60%) | 11 (41%) | 7 (54%) | 0.45 |
| Charlson Comorbidity Index (age adjusted), median (IQR) | 4 (0, 6) (n=15) | 3 (2, 5) (n=26) | 5 (2, 7) (n=11) | 0.64 |
| **Laboratory parameters** |  |  |  |  |
| CRP (mg/L), median (IQR) | 23.1 (18.3, 40.6) (n=15) | 117 (41.4, 193) (n=25) | 172 (110, 204) (n=13) | <0.01 |
| Haemoglobin (g/L), median (IQR) | 135 (116, 148) (n=15) | 139 (121, 142) (n=27) | 121 (111, 133) (n=13) | 0.27 |
| White cell count x10<sup>9</sup>/L, median (IQR) | 5.2 (4.3, 7.1) (n=15) | 6.2 (4.3, 8) (n=27) | 6.4 (4.2, 8.8) (n=13) | 0.38 |
| Lymphocytes x10<sup>9</sup>/L, median (IQR) | 1 (0.8, 1.5) (n=15) | 0.8 (0.5, 1) (n=27) | 0.7 (0.6, 0.8) (n=13) | <0.01 |
| D-dimer (ng/mL), median (IQR) | 599 (395, 972) (n=15) | 969 (503, 1386) (n=25) | 1215 (832, 1705) (n=13) | 0.22 |
| Ferritin (mcg/mL), median (IQR) | 230 (126, 448) (n=14) | 1084 (439, 1551) (n=25) | 525 (408, 631) (n=13) | <0.01 |
| LDH (units/L), median (IQR) | 262 (206, 269) (n=9) | 376.5 (337, 496.5) (n=16) | 330 (280, 525) (n=10) | 0.01 |
| **Treatment parameters** |  |  |  |  |
| Receipt of Antibiotics | 9 (60%) | 22 (81%) | 13 (100%) | 0.03 |
| Remdesivir | 3 (20%) | 9 (33%) | 4 (31%) | 0.65 |
| Dexamethasone | 6 (40%) | 18 (67%) | 9 (69%) | 0.18 |
| **Outcomes** |  |  |  |  |
| Admission to ICU | 3 (20%) | 13 (48%) | 6 (46%) | 0.18 |
| Requirement for > 24hr supplemental O2 | 5 (33%) | 21 (78%) | 13 (100%) | <0.01 |
| Mechanical ventilation | 0 (0%) | 7 (26%) | 2 (15%) | 0.09 |
| Duration of Antibiotics (IV+PO), days, median (IQR) | 4 (3, 5) (n=9) | 4 (2, 6) (n=23) | 4.5 (4, 8.5) (n=12) | 0.50 |
| Time to oral stepdown, days, median (IQR) | 2 (1, 3) (n=5) | 2 (1, 2) (n=13) | 3.5 (2.5, 4.5) (n=8) | 0.04 |

CRP, C-reactive protein; LDH, lactate dehydrogenase; IQR, interquartile range; O2, oxygen; IV, intravenous; PO, oral. \*Blood culture organisms: *Enterobacter cloacae*, PCT 38.8mcg/L; *Escherichia coli*, PCT 1.37mcg/L; *Staphylococcus epidermidis*, PCT 0.14mcg/L

PCT levels were significantly associated with antibiotic use (p=0.03). In total, 44/55 (80%) patients received antibiotic therapy during their admission. 9/15 (60%) patients with normal PCT received antibiotic therapy during their admission, compared with 22/27 (81%) patients in the medium PCT group, and 13/13 (100%) patients in the high PCT group. In those who received antibiotics, PCT was not associated with total duration of antibiotic therapy (p=0.50). PCT was associated with earlier de-escalation to oral therapy, with a median duration of 2 days prior to step down in the normal and medium PCT groups versus 3.5 days in the high PCT group (p=0.04).

PCT levels were associated with supplemental oxygen requirements during admission. 100% of patients in the high PCT group required supplemental oxygen, compared with only 35% of patients in the normal PCT group (p<0.01). PCT was not associated with requirement for ICU admission, or dexamethasone and remdesivir therapy. Serum PCT levels were associated with C-reactive protein (CRP) (p<0.01), lymphocytes (p=<0.01), ferritin (p<0.01) and lactate dehydrogenase (LDH) (p=0.01).

## Discussion

In our experience with PCT in COVID-19, we note that changes in serum PCT were associated with both initiation of antibiotic therapy, and intravenous-to-oral switch. These findings underline the potential utility of PCT as a component of antimicrobial stewardship interventions (10, 11). Indeed, all patients in the high PCT group received antibiotics during their admission, while 20% in the medium PCT group and 40% in the low PCT group did not receive any antibiotic therapy, suggesting clinicians were more comfortable withholding antibiotic therapy in patients with lower PCT; a potential stewardship intervention.

Once initiated, the total duration of antibiotic therapy (intravenous [IV] and oral) was the same across the groups. However, we found a significant association between PCT and de-escalation to oral therapy. Patients in the high PCT group received on average 1.5 days of additional IV antibiotics compared to those in the medium and low groups, suggesting increased clinician comfort in de-escalating antibiotics for patients without high PCT.

Whilst limited by small study size and non-randomized design, this study still suggests that, in COVID-19 patients, measurement of PCT, in conjunction with other clinical assessment, may have a role in prognostication and decision-making algorithms for a wider group of patients than only those admitted to ICU, aiding AMS interventions in this cohort.

## Data Availability

data is available by request to corresponding author

